# Genomic diversity and *BCL9L* mutational status in CTC pools predict overall survival in metastatic colorectal cancer

**DOI:** 10.1101/2024.12.26.24319649

**Authors:** Joao M. Alves, Nuria Estévez-Gómez, Roberto Piñeiro, Laura Muinelo-Romay, Patricia Mondelo-Macía, Mercedes Salgado, Agueda Iglesias-Gómez, Laura Codesido-Prada, Astrid DIez-Martín, Joaquin Cubiella, David Posada

**Author notes:** **Corresponding authors:** Joao M. Alves, David Posada. these authors contributed equally to this work (Joao M. Alves, Nuria Estévez-Gómez). **Competing interests:** The authors declare no competing interests.

## Abstract

The genomic diversity of circulating tumor cells (CTCs) and its clinical implications remain poorly understood. In this study, we characterized the mutational landscape of CTC pools stemming from 29 metastatic colorectal cancer (mCRC) patients and examined its relationship with disease progression. Our analysis revealed substantial variation in mutational burden among patients, with all CTC pools harboring non-silent mutations in key CRC driver genes. Importantly, higher genomic diversity in CTC pools was significantly associated with reduced overall survival. Furthermore, non-silent mutations in *BCL9L* emerged as a strong predictor of patient survival. Taken together, these findings underscore the potential of CTC genomic profiling as a promising prognostic tool in mCRC and highlight the need for further research into its clinical applications.

## 1. Introduction

Metastatic colorectal cancer (mCRC) remains one of the leading causes of cancer-related deaths worldwide (Morgan et al. 2023). Despite encouraging improvements in the detection and treatment of early-stage lesions (Zauber et al. 2012; Jodal et al. 2019), mCRC continues to have a dismal 5-year survival rate of roughly 14% (Shin, Giancotti, and Rustgi 2023), highlighting the urgent need for biomarkers that can effectively monitor disease progression, guide treatment decisions, and improve patient outcomes (Zygulska and Pierzchalski 2022; Oh and Joo 2020).

Multiple studies have now shown that tumors with higher levels of genomic diversity are associated with more aggressive disease (Joung et al. 2017), shorter recurrence times (Fernandez-Mateos et al. 2024), and poorer survival outcomes (McDonald et al. 2019). However, measuring genomic diversity from solid tissues often involves invasive and high-risk biopsy procedures. As an alternative, liquid biopsies from peripheral blood and other body fluids, such as saliva and urine, offer a minimally invasive procedure to measure tumoral genomic diversity through the analysis of circulating tumor DNA (ctDNA) and circulating tumor cells (CTCs) (Connal et al. 2023). Although most efforts in this regard have focused on the genomic characterization of ctDNA due to its relative ease of isolation (Calabuig-Fariñas et al. 2016), the analysis of CTCs provides a unique opportunity to study intact tumor cells, thus allowing for comprehensive whole-genome profiling of tumors (Lin et al. 2021). Moreover, CTCs are presumably responsible for metastatic seeding and can provide information for both primary and metastatic tumor development (Zhu et al. 2018; Castro-Giner and Aceto 2020).

In this context, CTC enumeration has already been validated as a reliable prognostic marker across several cancer types (Toss et al. 2014; Maly et al. 2019), including CRC (Miller, Doyle, and Terstappen 2010; Tsai et al. 2016). However, not all the CTCs within a patient are genetically identical, and initial sequencing studies have revealed that CTCs can exhibit significant genomic variation within patients (Mishima et al. 2017; Paoletti et al. 2018; Castro-Giner and Aceto 2020). The genomic diversity of a patient’s CTC pool may hold clinical significance for several reasons. First, the CTC pool could serve as a proxy for intratumoral genomic diversity, which, as mentioned above, has already been associated with increased cancer aggressiveness. Second, CTCs are considered precursors of metastasis. As such, a genomically diverse CTC population may increase the potential for successful dissemination, colonization, and growth in new environments (Ahmed and Gravel 2018). Nevertheless, the clinical significance of CTC genomic diversity remains largely unexplored.

In this study, we evaluated the clinical relevance of CTC genomic variation in mCRC. Using whole-exome sequencing data stemming from CTC pools collected from 29 mCRC patients, we provide a comprehensive description of the mutational landscape of CTCs and show that the overall genomic variation of CTC pools and the mutational status of the *BCL9L* gene can help predict patient survival.

## 2. Material & Methods

### 2.1 Clinical cohort and blood collection

We enrolled 25 mCRC patients diagnosed between October 2017 and January 2022 at the Hospital Universitario de Ourense, Spain, with histologically proven CRC. We collected a 15mL blood sample for each patient, stored in Transfix CTC-TVT tubes (Cytomark, UK) at room temperature. For patients undergoing chemotherapy treatment, blood samples were obtained just before a new chemotherapy cycle. All samples were obtained and collected after written informed consent from all subjects using a protocol approved by the Clinical Ethics Committee of Pontevedra-Vigo-Ourense (2018/301 approved 19/06/2018).

### 2.2 CTC enrichment and PBMC isolation

We processed all samples within 96 hours of collection using the Parsortix® platform (ANGLE plc, UK), which traps CTCs due to their larger size and lower compressibility than blood cells. For each patient, 10 mL of whole peripheral blood was loaded into a Parsortix microfluidic device. We enriched each sample in disposable Parsortix cassettes with a gap size of 6.5 µm (GEN3D6.5, Angle Inc., Guildford, UK) and at 99 mbar, according to the manufacturer’s guidelines. After separation, the captured cells were collected in 200 µL of PBS and stored at −80 °C.

To obtain the peripheral blood mononuclear cell (PBMC) fraction - to be used as “healthy controls” - we took the remaining 5 mL from each blood sample and performed Ficoll-Paque gradient centrifugation. We kept the PBMCs in RNAlater (Ambion, TX, USA) at -80 ºC until genomic DNA (gDNA) extraction.

### 2.3 Whole-genome amplification of CTC pools

Given the large collection volume (∼200 µl), we initially performed gDNA extraction of the CTC pools using the QIAamp DNA Blood Mini Kit (Qiagen, Germany) before performing whole-genome amplification (WGA) with the Ampli1™ kit (Menarini Silicon Biosystems, Italy). We carried out the WGA starting with 1 µl of gDNA and included positive (10 ng/µl REPLIg human control kit, Qiagen, Germany) and negative controls (DNase/RNase free water). We worked in a laminar-flow hood to avoid contamination and used a dedicated set of pipettes and UV-irradiated plastic materials. We evaluated the quality of the amplified product with the Ampli1 QC Kit (Menarini Silicon Biosystems, Italy), a PCR-based assay to establish DNA integrity. Samples that produced at least two of the four expected PCR fragments were selected for the following steps: increasing the total double-stranded DNA (dsDNA) with the Ampli1 ReAmp/ds kit (Menarini Silicon Biosystems, Italy) and WGA adaptor removal. The latter was carried out by incubating a mixture of 5 µl of NEBuffer 4 10X (New England Biolabs, MA, USA), 1 µl of MseI 50U/µl (New England Biolabs, MA, USA), 19 µl of nuclease-free water and 25 µl of dsDNA at 37 ºC for 3 h, with a final step of enzyme inactivation at 65 ºC for 20 min. Finally, we purified the samples with 1.8X AMPure XP beads (Agencourt, Beckman Coulter, CA, USA), quantified the DNA yield with Qubit 3.0 fluorometer (Thermo Fisher Scientific, MA, USA), and checked the amplicon size distribution with the D1000 ScreenTape System in a 2200 TapeStation platform (Agilent Technologies, CA, USA).

### 2.4 PBMCs gDNA isolation

In parallel, we used the QIAamp DNA Blood Mini Kit (Qiagen, Germany) to extract gDNA from the PBMCs. We estimated DNA yield using the Qubit 3.0 fluorometer (Thermo Fisher Scientific, MA, USA) and DNA integrity with the Genomic DNA ScreenTape Assay (Agilent Technologies, CA, USA).

### 2.5 Whole-exome sequencing

Sequencing libraries were constructed at the Spanish National Center for Genomic Analysis (CNAG; http://www.cnag.crg.eu) with the SureSelect XT and Agilent Human Exon v5 kits (Agilent Technologies, CA, USA). In total, 25 amplified CTC pools and 25 PBMCs gDNA samples were whole-exome sequenced (WES) at 100X and 60X, respectively, on an Illumina NovaSeq 6000 (PE100) at CNAG. In addition, we also included matched CTC pools and PBMCs WES datasets previously generated in our lab from four mCRC patients (“PP”) (Alves et al. 2022) (Sequence Read Archive accession code PRJNA886718).

### 2.6 Data processing

After trimming amplification and sequencing adapters from the raw FASTQ files, we aligned the sequencing reads to the Genome Reference Consortium Human Build 37 (GRCh37) using the MEM algorithm in the BWA software (Li 2013). Following the GATK’s standardized best-practices pipeline (Van der Auwera et al. 2013), we filtered out reads with low mapping quality, performed a local realignment around indels, and removed PCR duplicates.

### 2.7 Somatic variant calling

We identified somatic single nucleotide variants (SNVs) and short insertions and deletions (indels) for each patient using the paired-sample variant-calling approach implemented in MuTect2 software (Benjamin et al. 2019) (i.e., CTC pools (tumor) + PBMCs (healthy control)). We then applied GATK FilterMutectCalls to remove calls in problematic sequence contexts (“--orientation-bias-artifact-priors”) or due to potential cross-sample contamination (“--contamination”). Genotypes supported by < 10 total reads, with < 2 alternative reads, or with a variant allele frequency (VAF) ≤ 0.075 were set as missing, and variants composed only of missing genotypes were removed. We annotated the variants with Annovar (v.20200608) (Wang, Li, and Hakonarson 2010). In addition, and for each patient, we estimated somatic copy number gains, losses, loss-of-heterozygosity (LOH) events, tumor purity, and global ploidy status with Sequenza (Favero et al. 2015) under default settings.

### 2.8 MSI status

Furthermore, we identified the microsatellite stability status of each patient with MSIsensor-pro (Jia et al. 2020), following the recommended ‘best practices’ for paired-sampling analysis. We classified patients with more than 30% of microsatellites mutated as microsatellite instable (MSI), while all other patients were categorized as microsatellite stable (MSS), as in Heide *et al*. (2022).

### 2.9 Mutational signatures

We ran the SIGNAL web tool (https://signal.mutationalsignatures.com/) (Degasperi et al. 2020) under default parameters to identify the single base substitution (SBS) signatures active in each patient (Alexandrov et al. 2020). As recommended by the authors, SBS fitting was performed using candidate SBS signatures from CRC.

### 2.10 Estimation of genomic diversity in CTC pools

The genomic diversity of a CTC pool can be measured in multiple ways depending on the type and features of the variants selected. Here, we computed 15 diversity metrics, including mutational burden (MB), the proportion of aberrant genome (PAG) (Schumacher et al. 2017), the mutant allele tumor heterogeneity score (MATH) (Mroz and Rocco 2013), number of SNVs and indels at CRC driver genes, number of copy number alterations (CNAs) at CRC driver genes, and mutation status at recurrently mutated CRC driver genes. Since our cohort only included one MSI patient, all downstream analyses were restricted to the MSS cases (N=28).

#### 2.10.1 Mutational burden (MB)

For each patient, we measured the CTC pool mutational burden (MB) as the total number of SNVs that passed our filtering thresholds, normalized by the size of the exome captured. We report MB in units of number of SNVs / megabase.

#### 2.10.2 Proportion of aberrant genome (PAG)

We also measured, for each patient, the proportion of the autosomal genome affected by copy number gains, losses, and LOH events.

#### 2.10.3 Mutant allele tumor heterogeneity (MATH)

We estimated for each patient the CTC-pool mutant-allele tumor heterogeneity (MATH) score (Mroz and Rocco 2013), which is based on the distribution of SNV and indel allelic frequencies and calculated as:

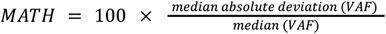

Where VAF is the variant allele frequency of SNVs and indels. Since MATH scores are sensitive to unreliable VAF at sites with insufficient sequencing depth (Makrooni, O’Sullivan, and Seoighe 2022), we only considered SNVs and indels with ≥ 25 total reads and ≥ 5 alternative reads.

#### 2.10.4 Number of SNVs and indels at CRC driver genes (CRC-mut)

We additionally measured the total number of the non-silent SNVs and indels (i.e., mutations that alter the amino acid sequence of a protein and may affect its function) overlapping with CRC driver genes at the IntOGen portal (release 2023) (Gonzalez-Perez et al. 2013).

#### 2.10.5 Number of CNAs at CRC driver genes (CRC-CNA)

We estimated the number of copy number alterations (CNAs), which includes gains, losses, and LOH events in the CTC pools overlapping with CRC driver genes at IntOGen (release 2023).

#### 2.10.6 Mutation status at CRC driver genes

For CRC driver genes with recurrent non-silent SNVs/Indels in more than four patients, we additionally considered their mutation status (i.e., unmutated vs. mutated) as a diversity metric.

### 2.11 Correlation of CTC genomic diversity with overall survival

We correlated the above diversity metrics with overall survival (OS). To select which metrics should be included in a multivariate Cox proportional hazards (CPH) analysis, we assessed them individually in a univariate CPH analysis. We used the raw values for the continuous metrics. For the discrete ones (i.e., mutation status and gender), we used “unmutated” and “female” as reference groups, respectively.

Additionally, we created a binary version of each continuous metric (i.e., bMB, bAge, bLoT, bPAG, bMATH, bCRC-mut, and bCRC-CNA) for inclusion in a univariate analysis by splitting the patients into two groups given an optimal threshold. This threshold was determined using CutoffFinder (Budczies et al. 2012), which fits a CPH model to the explanatory (diversity metric) and response (OS) variables, identifying the optimal cutoff as the value that provides the most statistically significant split based on a log-rank test (**Table S1**).

Adopting a similar strategy to Fernandez-Mateos *et al*. (2024), we only included diversity metrics with a p-value ≤ 0.1 in the multivariate analysis. Importantly, when the continuous and binary versions of a metric returned p-values ≤ 0.1, only the continuous version was included in the multivariate analysis. All CPH analyses were performed using the survival (Therneau 2001) and survminer (Kassambara, Kosinski, and Biecek 2016) R packages.

### 2.12 Evaluating custom and commercial cancer gene panels for CTC genomic diversity estimation

To identify a cost-effective strategy for the genomic characterization of CTC pools, we constructed and evaluated two customized gene panels, each consisting of a subset of the most frequently mutated genes in our cohort. These panels were tested to determine their ability to provide diversity estimates comparable to those derived from WES. The first panel, CRC-P22, comprised 22 genes harboring > 15 mutations across patients. The second panel, CRC-P55, included 55 genes with > 10 mutations across patients. In addition to these custom panels, we assessed the performance of two commercially available NGS panels, the Pillar® oncoReveal Solid Tumor v2 Panel (48 genes; Pillar Biosciences, MA, USA) and the Oncomine Precision Assay (50 genes; Thermo Fisher Scientific, MA, USA). A list of the genes from the four panels is provided in **Table S2**.

We extracted the SNVs and indels for each panel and calculated the MATH scores for each patient as in Section 2.10.3. Importantly, given the much smaller number of variants in these panels, we did not apply any filter based on read counts, as previously done. Finally, we assessed the clinical relevance of these panels by correlating their derived MATH scores with OS using a univariate CPH analysis. As before, we additionally tested a binary MATH (bMATH) version by dichotomizing patients at optimal threshold values determined using CutoffFinder.

### 2.13 Declaration of AI-assisted copy editing

During the preparation of this work we have used ChatGPT 4.0 turbo to improve language and readability. After using this tool/service, we reviewed and edited the content as needed and took full responsibility for the content of the publication.

## 3. Results

### 3.1 Genomic characterization of CTC pools from the mCRC cohort

A brief overview of the cohort is shown in **Fig. 1A**. The comprehensive clinical data for this cohort, including tumor and treatment details, is available in **Table S3**. Ten (34.5%) patients were female, and 19 (65.5%) were male, with a mean age at the time of mCRC diagnosis of 66.28 ± 16.33 years.

**Figure 1.**
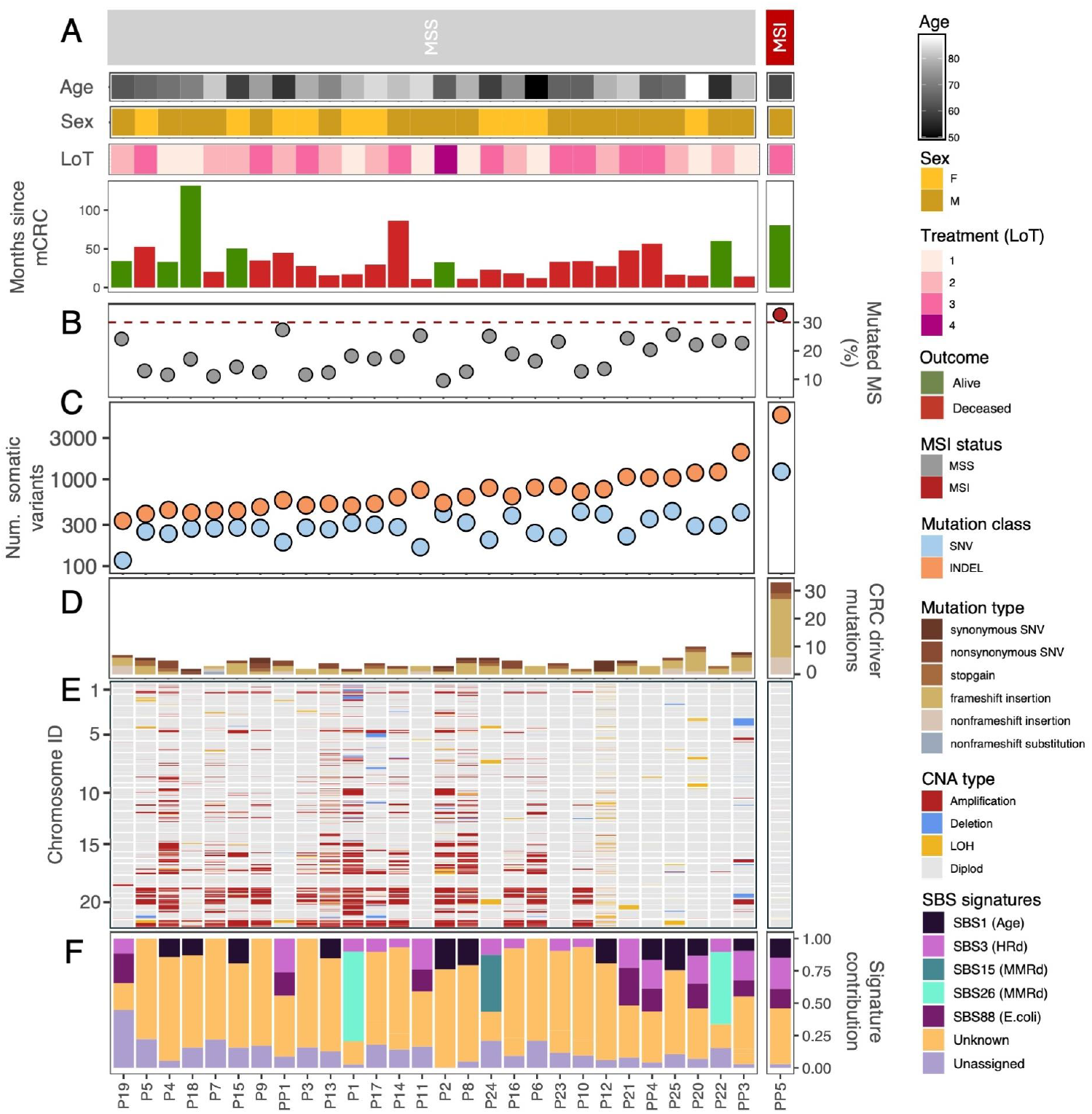
Clinical and genomic characterization of mCRC cohort. mCRC patients are split based on MS status and ordered according to their total number of somatic variants. **A**. Age, sex, number of lines of treatment (LoT), and months since mCRC diagnosis. **B**. Proportion of mutated microsatellites. Data points are colored by the MS status (grey = “MSS”, red = “MSI”). The dashed red line depicts the threshold for MSI classification (i.e., 30% mutated microsatellites). **C**. Total number of somatic mutations identified with MuTect2, including SNVs (light blue) and indels (orange). **D**. Total number of mutations affecting known CRC driver genes in each patient. Bars are colored according to the mutation type. **E**. Genome-wide copy number profiles for each patient. Only autosomes are shown, with chromosomes ordered from 1 to 22. Genomic regions are colored according to CNA type. **F**. Contribution of the different mutational signatures. HRd = homologous recombination deficiency; MMRd = mismatch repair deficiency. SBS signatures with unknown etiology - SBS8, SBS93, SBS97, SBS121, and SBS123 - were collapsed into a single “Unknown” category.

After sequencing, we identified one patient (PP5) exhibiting microsatellite instability (MSI) (**Fig. 1B**). As expected, this patient displayed a substantially higher SNV and indel burden (**Fig. 1C**). Specifically, for PP5, we called a total of 1,224 SNVs and 5,509 indels. In contrast, we estimated 116-429 SNVs and 331-2,043 indels for the other patients.

We identified a large number of mutations in known CRC driver genes, including non-synonymous changes and stop codon gains in *APC, KRAS*, and *PIK3CA*. Notably, the number of mutations in CRC driver genes varied significantly within the cohort, ranging from two mutations in patients P1, P3, P10, and P18 (MSS) to 33 mutations in patient PP5 (MSI) (**Fig. 1D**). We also inferred a substantial number of CNAs, with considerable differences among patients (**Fig. 1E**). Consistent with previous reports (Heide et al. 2022), the MSI patient PP5 exhibited a predominantly diploid copy number profile.

We found ten single-base substitution signatures, including signatures commonly observed in CRC, such as SBS1 (age-related), SBS3 (homologous recombination deficiency) and SBS88 (associated with exposure to *E*.*coli* bacteria) (**Fig. 1F**). Nevertheless, there was a significant contribution of signatures of unknown etiology - SBS8, SBS93, SBS97, SBS121 and SBS123 - all of which are relatively infrequent in CRC.

### 3.2 CTC pools show high levels of genomic variation

We observed high levels of genomic variation in the CTC pools (**Fig. 2A**), with substantial variation among patients. The MB estimates ranged from 0.95 to 2.89 SNVs/Mb, PAG values from 0.05% to 40.5%, and MATH scores between 32.38 and 67.63.

**Figure 2.**
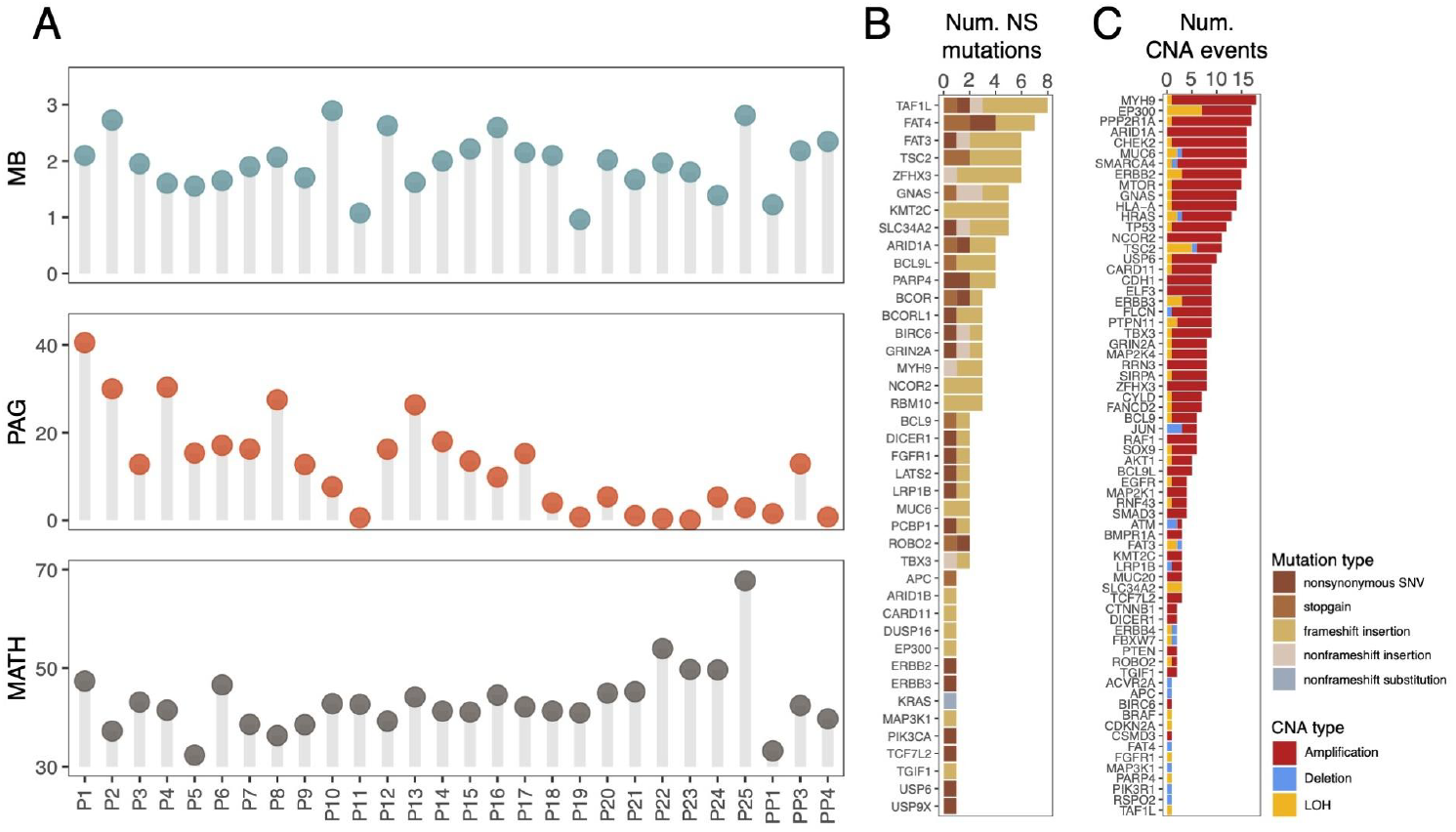
Genomic diversity in CTC pools. **A**. Lollypop plots depict the mutational burden (MB), the proportion of aberrant genome (PAG), and the mutant allele tumor heterogeneity (MATH) score for each patient. **B**. Number of non-silent (NS) SNVs and indels affecting known CRC driver genes. Bars are colored according to mutation type. **C**. Number of CNAs overlapping known CRC driver genes. Bars are colored according to CNA type.

In addition to the substantial variability in the number of non-silent SNVs in CRC driver genes observed among patients (**Fig. 1D**), we also identified several genes that were recurrently mutated across multiple patients (**Fig. 2B**). In particular, ten driver genes were independently mutated in at least four patients, including *ARID1A* (N=4), *BCL9L* (N=4), *BIRC6* (N=5), *FAT3* (N=5), *FAT4* (N=5), *KMT2C* (N=5), *PARP4* (N=4), *TAF1L* (N=7), *TSC2* (N=4), and *ZFHX3* (N=5). Similarly, CNAs encompassing CRC drivers were also widespread (**Fig. 2C**), with *ARID1A* (N=16), *MYH9* (N=17), and *PPP2R1A* (N=16) having the highest frequency of copy gains, while *EP300* (N=7) and *TSC2* (N=6) had the highest number of deletions and LOH events.

### 3.3 MATH score and *BCL9L* status predict survival outcome in mCRC

The univariate analyses indicated that patient age is significantly associated with shorter OS, both when analyzed as a continuous variable (Hazard Ratio [HR]=1.07, 95% CI: 1.02–1.12, p=0.00894) and when dichotomized (HR=3.67, 95% CI: 1.39–9.67, p=0.00859) (**Fig. 3A**). Patients in the High-bMATH group (HR=3.45, 95% CI: 1.36–8.75, p=0.00895) also had poorer OS compared to those in the corresponding “Low” groups.

**Figure 3.**
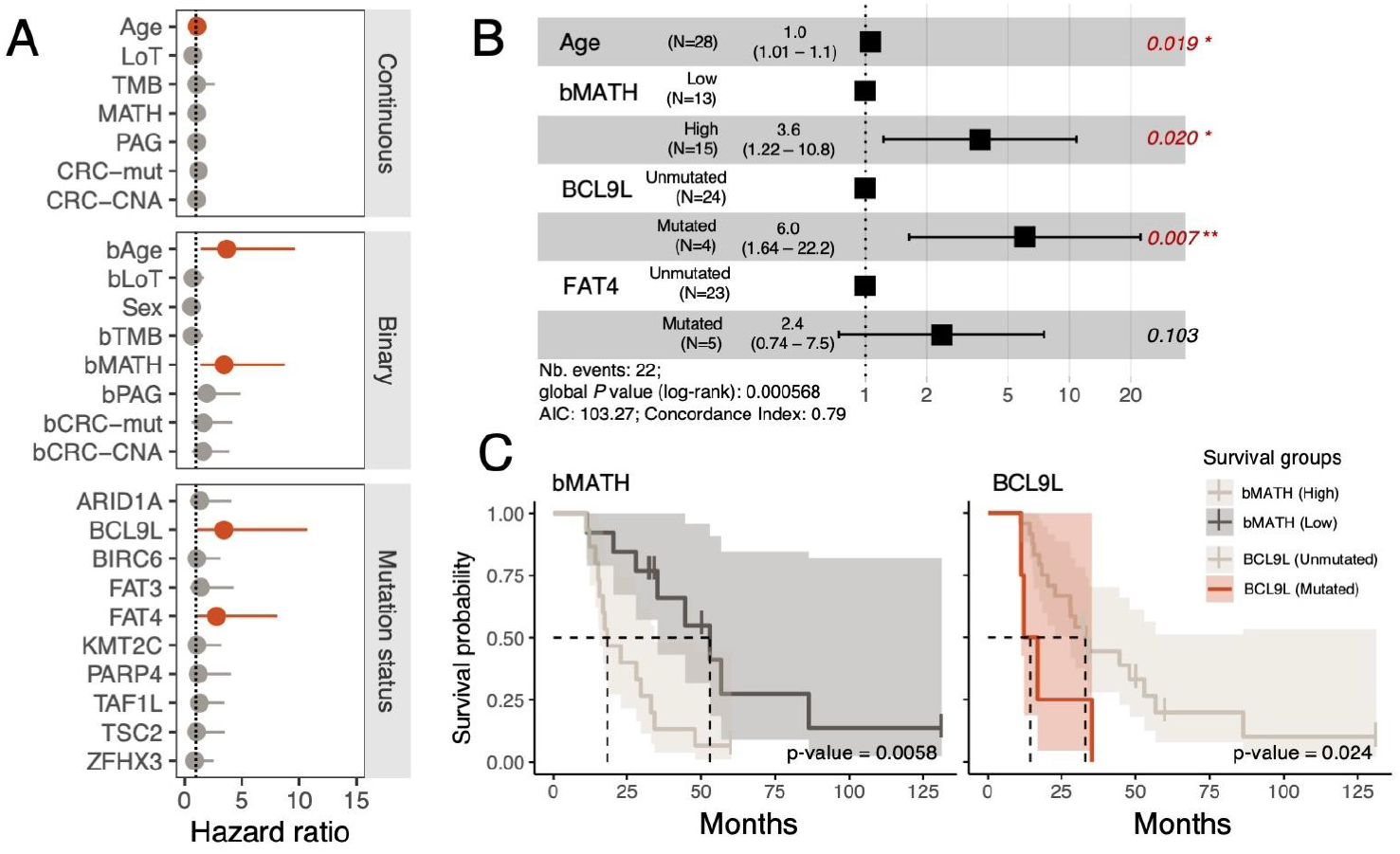
Impact of CTC genomic diversity on overall survival. **A**. Univariate CPH analysis for OS using all diversity metrics. Dots are the point estimate of the Hazard ratio, while the lines represent the 95% confidence interval (CI). The “Low” group was used as a reference in the binary version of all continuous variables. Hazard ratios with p-values < 0.1 are colored in red. **B**. Multivariate CPH analysis of survival time for the significant metrics and covariates in the univariate CPH analysis shown in A. The forest plot shows a 95% CI of HRs and the covariate p-values. Significant p-values are colored in red. **C**. Kaplan–Meier survival curves and p-values for bMATH and *BCL9L* on the mCRC cohort. The shaded area around each survival curve depicts the 95% CI.

Notably, a subgroup of patients (n=4) with non-silent mutations in *BCL9L* also displayed significantly worse OS (HR=3.71, 95% CI: 1.18–11.7, p=0.02). Importantly, *BCL9L* has been linked to increased genomic heterogeneity by promoting aneuploidy tolerance in CRC (López-García et al. 2017). In our cohort, patients with *BCL9L* mutations exhibited a higher fraction of CNAs, as reflected by elevated PAG estimates (**Fig. S1A**), but this did not translate into higher MATH scores (**Fig. S1B**). Similarly, patients (n=5) harboring non-silent mutations in *FAT4* displayed significantly worse OS (HR=2.80, 95% CI: 0.966–8.09, p=0.058).

A multivariate analysis including patient age, bMATH, as well as *BCL9L* and *FAT4* mutation status, confirmed that both high bMATH and mutated *BLC9L* status were significant prognostic factors, with HRs of 3.6 (95% CI=1.22–10.8, p-value=0.020) and 6.0 (95% CI=1.64–22.2, p-value=0.007), respectively (**Fig. 3B**). Accordingly, the survival curves indicate that patients in the Low-bMATH group survive much longer (median survival time: 52.9 months) than their High-bMATH counterparts (median survival time: 18.3 months) (**Fig. 3C**). Similarly, patients carrying *BCL9L* mutations experienced a notable reduction in OS (median survival time: 14.5 months) compared to those without them (median survival time: 33.1 months).

### 3.4 CTC diversity from gene panels can also predict patient survival

To explore a practical and low-cost alternative for CTC genomic profiling, we designed two gene panels – CRC-P22 and CRC-P55 – each comprising the most frequently mutated genes in our cohort (**Fig. 4A**). The MATH scores derived from the CRC-P55 panel were highly correlated with those obtained from the WES data, whereas the CRC-P22 panel showed weaker concordance (**Fig. 4B-C)**. Importantly, MATH estimates from the CRC-P55 panel were significantly associated with worse OS (**Fig. 4D**), both as a continuous variable (HR = 1.04, 95% CI: 1.01–1.07, p = 0.0223) and as a binary variable (HR = 4.83, 95% CI: 1.66–14.0, p = 0.00377). Patients in the high-bMATH group exhibited a significantly shorter median OS (20.4 months) compared to those in the low-bMATH group (52.9 months) (**Fig. 4E**). On the other hand, neither of the commercial panels showed a significant association between MATH scores and OS (**Fig. S2**).

**Figure 4.**
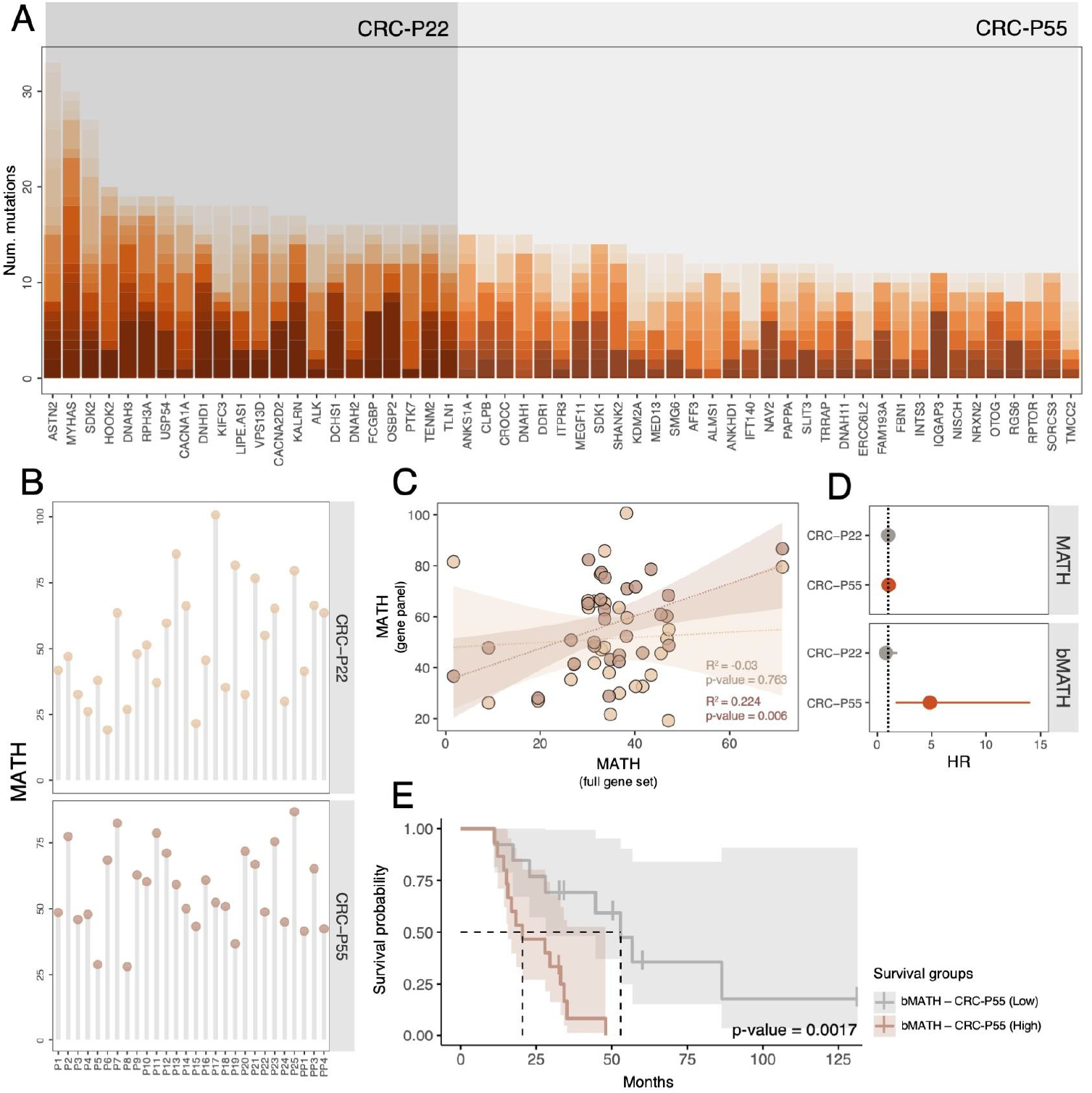
Optimized gene panel analysis. **A**. Genes included in the CRC-P22 and CRC-P55 panels were sorted according to mutation counts. Different color shades represent the different patients. **B**. MATH scores for each panel and patient. **C**. Scatter plot describing the similarity of MATH scores between gene panels and WES. Solid lines represent the best fit from regression analysis. R^2^ scores and *p-values* are shown on the bottom right side of the plot. Lighter and darker shades of brown highlight the MATH scores from CRC-P22 and CRC-P55, respectively. **D**. Univariate CPH analysis of survival time and MATH and bMATH for both gene panels. Dots are the Hazard ratio (HR) point estimate, while the lines represent the 95% CI. In the bMATH, the “Low” group was used as a reference. P-values < 0.05 are colored in red. **E**. Kaplan–Meier survival curves and p-values for bMATH of CRC-P55 gene panel. The shaded area around each survival curve depicts the 95% CI.

## 4. Discussion

The prognostic value of CTC enumeration has already been established, with higher CTC counts linked to aggressive disease across multiple cancer types, including CRC (Tsai et al. 2016). However, CTC populations can be genomically heterogeneous (Alves et al. 2022), a factor that, in the case of primary tumors, has already been linked to poor prognosis (Joung et al. 2017; Fernandez-Mateos et al. 2024; McDonald et al. 2019).

Here, we found that, in mCRC, CTC populations with higher MATH scores - a measure of genomic diversity - or with non-silent mutations in *BCL9L* are significantly associated with poorer overall survival. To our knowledge, this is the first study showing an association between CTC genomic diversity and cancer outcome. Intuitively, larger overall genetic variation, as measured by the MATH scores, likely reflects the presence of distinct subclones, some of which could proliferate more rapidly, have a higher propensity for metastasis, or better evade cancer drugs (McGranahan and Swanton, 2017). In this context, genomic diversity estimates derived from CTC pools may not only serve as prognostic biomarkers but might also contribute to real-time monitoring of disease progression (through longitudinal sampling). Increasing CTC heterogeneity over time could signal the emergence of resistant clones, potentially allowing for earlier clinical intervention. Conversely, a decline in diversity following treatment might suggest a favorable response.

Since the MATH scores in this study were derived from whole-exome sequencing - which may be impractical in clinical settings due to high costs and complex bioinformatics workflows (Bertier, Hétu, and Joly 2016) - we explored the potential of smaller gene panels as an alternative. Our findings revealed a strong correlation between MATH scores derived from a 55-gene panel and survival, offering a cost-effective strategy for clinical diagnostics and patient stratification.

Importantly, we also observed that non-silent mutations in *BCL9L* were associated with reduced survival. *BCL9L* has been recently identified as a driver of aneuploidy tolerance in CRC, thereby increasing genomic instability (López-García et al. 2017), which has already been proposed as a prognostic marker in several cancer types (Hosea et al. 2024).

Despite these promising findings, the relatively small sample size of our cohort may limit the generalizability of our conclusions, and larger cohorts will be needed to fully validate the relationship between CTC genomic data and patient outcomes.

In summary, our study demonstrates for the first time a relationship between CTC genomic diversity and overall survival in mCRC, underscoring the potential of genomic profiling of CTCs as a novel prognostic tool in mCRC and highlighting the need for further research into the clinical applications of CTC genomics, which could ultimately improve personalized treatment strategies for cancer patients.

## Data availability

Raw exome sequencing data from CTC pools, together with matching healthy samples, have been deposited in the Sequence Read Archive database under the accession code ########. All data supporting the findings of this study are available within the article and its supplementary information files.

## Author contributions

D.P. and J.M.A. conceived the study, designed the analyses, and obtained the funding to perform the experiments. M.S. and J.C. supervised sample collection and obtained patient information. A.I.-G., L.C.-P., and A.D.-M. collected the blood samples. R.P., L.M.R., and P.M. performed the CTC enrichment experiments.

N.E.G. performed whole-genome amplification and obtained DNA from CTC pools and healthy PBMCs.

J.M.A. performed the analyses. All authors read and approved the final manuscript.

## Competing interests

The authors declare that they have no competing interests.

## Acknowledgments

We thank the Supercomputation Center of Galicia (CESGA; https://www.cesga.es) for providing all the computational resources needed for this study. This work was supported by the Spanish Ministry of Science and Innovation - MICINN (PID2019-106247GB-I00 awarded to DP) and by an AXA Research Fund postdoctoral grant (awarded to J.M.A.). DP receives further support from the Galician government (ED431C 2022/26). JMA is currently supported by an AECC-Investigator grant (INVES20007FERN). R.P. is currently supported by an AECC-Investigator grant (INVES234992PIÑE). L.M.R. is supported by a contract “Miguel Servet” from ISCIII (CP20/00119).

## Notes

### Competing Interest Statement

The authors have declared no competing interest.

### Author Declarations

All samples were obtained and collected after written informed consent from all subjects using a protocol approved by the Clinical Ethics Committee of Pontevedra-Vigo-Ourense (2018/301 approved 19/06/2018). This study was approved by the Clinical Ethics Committee of Pontevedra-Vigo-Ourense

### Summary of Updates

In this version, we have updated the analysis by including additional clinical variables (lines of treatment) and the latest clinical survival details.

