## Supplementary figures for "Genomic diversity and *BCL9L* mutational status in CTC pools predict overall survival in metastatic colorectal cancer"

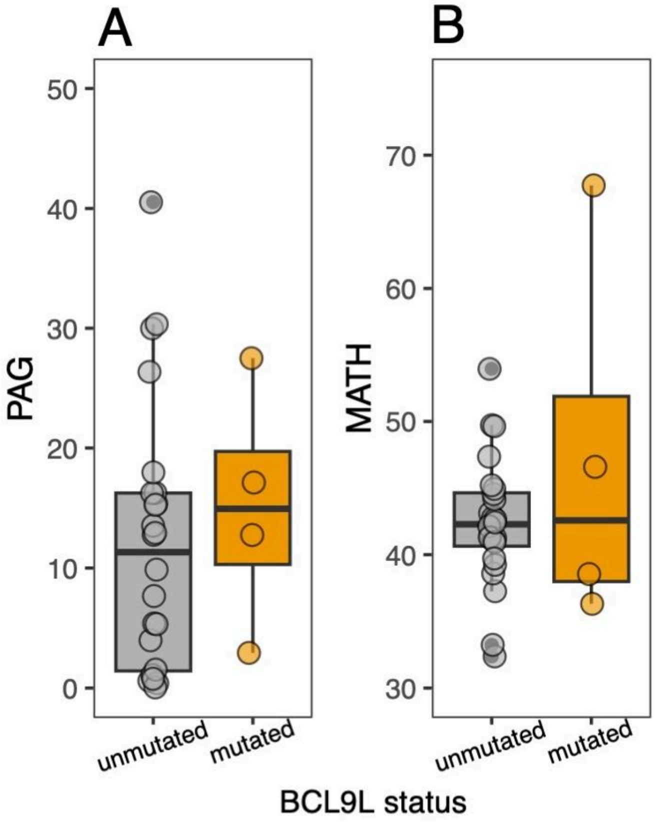


**Figure S1. PAG and MATH scores according to *BCL9L* status. A.** PAG scores for patients with unmutated (grey) and mutated (orange) *BCL9L* gene. **B.** MATH scores for patients with unmutated (grey) and mutated (orange) *BCL9L* gene.

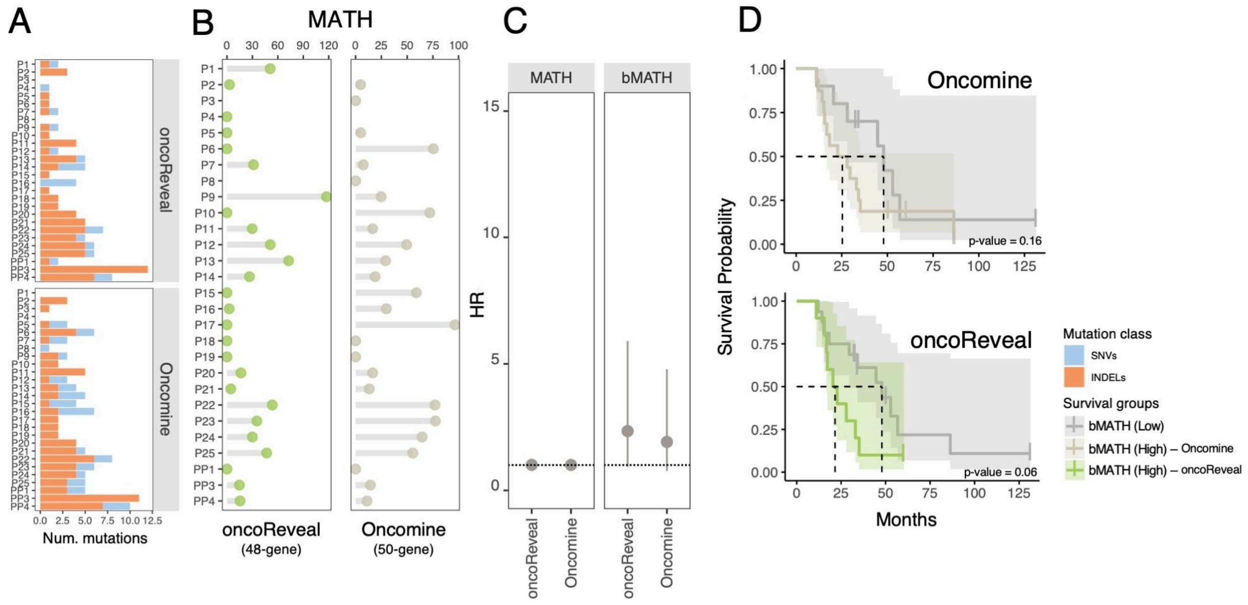


**Figure S2. Commercial NGS panel analysis. A.** Mutation counts in each gene panel, including SNVs (light blue) and indels (orange). **B.** MATH scores for each panel and patient. **C.** Univariate CPH analysis of survival time and MATH and bMATH for both gene panels. Dots are the point estimate of the Hazard ratio (HR), while the lines represent the 95% CI. In the bMATH, the "Low" group was used as a reference. **D.** Kaplan–Meier survival curves and p-values for the bMATH score of both gene panels. The shaded area around each survival curve depicts the 95% CI.
